# Contrast-Enhanced Magnetic Resonance Imaging Based T1-Mapping and Extracellular Volume Fractions are Associated with Peripheral Artery Disease

**DOI:** 10.1101/2023.03.03.23286787

**Authors:** Asem Fitian, Michael C. Shieh, Olga A. Gimnich, Tatiana Belousova, Addison A. Taylor, Christie M. Ballantyne, Jean Bismuth, Dipan J. Shah, Gerd Brunner

**Author notes:** Address for correspondence: Gerd Brunner, MS, PhD, 500 University Drive H047, Hershey, PA 17033. These authors contributed equally (Asem Fitian, Michael C. Shieh).

## Abstract

**Background:** Extracellular volume fraction (ECV), measured with contrast-enhanced magnetic resonance imaging (CE-MRI), has been utilized to study myocardial fibrosis but its role in peripheral artery disease (PAD) remains unknown. We hypothesized that T1-mapping and ECV differ between PAD patients and matched-controls.

**Methods and Results:** A total of 37 individuals (18 PAD, 19 controls) underwent 3.0T CE-MRI at the mid-calf. T1-mapping was done before and after gadolinium contrast with a motion corrected Modified Look Locker Inversion Recovery (MOLLI) pulse sequence. T1 values were calculated with a 3-parameter Levenberg-Marquardt curve fitting algorithm. T1-maps were quantified in 5 calf muscle compartments (anterior [AM], lateral [LM], and deep posterior [DM] muscle groups; soleus [SM], and gastrocnemius [GM] muscles). Averaged peak blood pool T1 values were obtained from the posterior (PT) and anterior tibialis (AT); and peroneal artery (PE). PAD ECV was calculated for each muscle group. T1 values and ECV are heterogeneous across calf muscle compartments. Native peak T1 values of the AM, LM, and DM were significantly higher in PAD patients compared to controls (all p<0.028). ECV of the AM and SM were significantly higher in PAD patients compared to controls (AM: 26.4% (21.2, 31.6) vs. 17.3% (10.2, 25.1), p=0.046; SM: 22.7% (19.5, 27.8) vs. 13.8% (10.2, 19.1), p=0.020).

**Conclusions:** Native peak T1 values and ECV fractions of several calf muscle compartments are higher in PAD patients compared with matched-controls. These data suggest a potential utility of ECV to noninvasively determine skeletal muscle fibrosis in PAD.

## Introduction

A natural outgrowth of the past decade’s advancements in magnetic resonance imaging (MRI) and computed tomography (CT) has been the enhanced diagnostic accuracy of myriad ailments owing to the more robust processing power of state-of-the-art imaging modalities^1^. Notwithstanding the reproducibly more accurate detection of pathologies previously encumbered by limitations in imaging quality and efficiency, paradigm-changing advances in the disease monitoring and diagnosis realms—ones that detect diseases during their clinically silent stages rather than the conventional presentation when curative interventions are at best a long shot— continue to elude clinical fruition.

With advances in resolution and processing power occurring amid a deep learning and artificial intelligence (AI) revolution^2^, the opportunity has emerged to, at still-more granular radiographic levels, define pathophysiologic underpinnings that not only more accurately delineate whether an anomaly is present but stratify the abnormality based on the most probable culprit central process or disease^3,4^. Such advancements in precision medicine and radiography have, for example, already been realized in the breast cancer diagnosis realm with significantly improved early detection of what would otherwise, in the setting of conventional surveillance modalities, prove clandestine tumors—and with superior efficacy capable of surmounting historically challenging clinical diagnostic conundrums^5,6^.

To this end, radiomics-driven solutions are continually demonstrating game-changing potential in the effort to deliver early diagnostic and disease monitoring solutions to the cardiovascular disease arena down to previously unseen pathophysiologic levels. Notable advancements to this end include Bayesian networks that leverage data from stress echocardiography and CT angiography (CTA) to accurately predict future cardiovascular events such as myocardial infarction (MI) or death^7,8^ a machine learning algorithm that outperforms conventional diagnostic strategies for accurate detection of heart failure with preserved ejection fraction^9^ among myriad other novel algorithms that show promise in the diagnosis of obstructive coronary stenoses^10^, myocardial infarction^11^, and peripheral vascular disease (PVD)^12^. Owing to the grim 10-year mortality rate facing patients with limb claudication that eclipses 50% when considering age and comorbid factors^13^ and the increased rate of myocardial infarction, stroke, and cardiovascular death in patients with both symptomatic and asymptomatic peripheral artery disease (PAD)^14^, it is imperative that one such paradigm change occurs in the surveillance for the earliest warning signs of limb ischemia.

One avenue of promise has emerged and stems from advances in quantifying the extracellular volume fraction (ECV) in the myocardium measured with contrast-enhanced magnetic resonance imaging (CE-MRI). The T1 relaxation time, a measure of how fast the nuclear spin magnetization equilibrates after a radiofrequency pulse in MRI, is an important tool for highlighting soft-tissue contrast. T1 times are specific to and span myriad pathologies, from fibrosis to fatty infiltration. Owing to the pathological underpinnings of fibrosis, arising from derailments in and expansion of extracellular matrix (ECM) remodeling^15^, contrast-enhanced quantification of ECV has proven a useful means for detecting microscopic areas of ischemia, myocardial scarring, and perfusion^13,16^.

Advances in cardiac MRI leverage pre- and post-contrast injection in T1 sequences to disclose precise, focal areas of pathology by measuring the rate of change of T1 relaxation relative to the degree of contrast pooling, with T1 times corresponding to pathologic states. For example, acute membrane rupture emblematic of acute MI and myocarditis allows aberrant infiltration of gadolinium contrast into myocytes, resulting in increased gadolinium concentration, prolonged T1 relaxation times, and the corresponding hyper-enhancement hallmark of acute MI. By the same token, in chronic MI, scarring prevails, with a corresponding expansion of the interstitium that leads to increased gadolinium concentration, hyper-enhancement, and correspondingly prolonged T1 times.

Previous works^17 18^ have demonstrated the potential reproducibility of T1 mapping beyond central cardiovascular to peripheral vascular pathophysiologic areas^19^. In this study, we investigated T1 times in PAD patients and matched controls. We hypothesized that differences in T1 relaxation times will be observed in PAD patients compared with appropriately matched controls.

## Methods

The study was approved by the institutional review board and conducted in keeping with the Helsinki Declaration of 1973. Study participants provided informed consent, and the study obtained approval from the institutional review boards at the Houston Methodist Hospital and the Michael E. DeBakey Veterans Affairs Medical Center, Houston, TX. Suitable patients from the Houston Methodist Hospital and the Michael E. DeBakey Veterans Affairs Medical Center in Houston, TX were identified using inclusion and exclusion criteria in the medical record. PAD patients and controls without PAD were recruited and matched on the basis of demographic and clinical characteristics. Standard of care continued uninterrupted throughout study subject participation in this observational imaging work. Patients with contraindications to MRI and those with an estimated glomerular filtration rate (eGFR) ≤40 mL/min/1.73.m^2^ were excluded from this study.

Participants were positioned on the MRI table feet first in the supine position, and imaging was performed at the mid-calf level. Efforts to minimize motion artifact were employed using a bilateral leg coil employed during the imaging runtime. A gadolinium-based contrast agent was administered intravenously (gadopentetate dimeglumine [Magnevist, Bayer Healthcare, Whippany, New Jersey, USA at 0.2 mmol/kg, or gadobutrol [Gadavist, Bayer Healthcare, at 0.1 mmol/kg) with flow rates of 2-4 ml/sec proceeded by a 20 ml saline flush. MRI scans were saved in the DICOM format prior to rendering and further analysis. The arterial lumens of the anterior tibialis (AT), posterior tibial (PT), and peroneal artery (PE) were segmented and visualized as permitted by contrast enhancement using Sante DICOM Editor Version 3.0 (Santesoft, Nicosia, Cyprus).

The reproducibility and quality of the lumen frame analysis and interpretation were assessed through inter- and intraobserver reproducibility analyses. Cross-sectional leg muscle area (CSLMA) was measured using our previously described methodology.^18^

T1-mapping was done before and after gadolinium contrast administration with a motion-corrected Modified Look-Locker Inversion Recovery (MOLLI) pulse sequence. T1 values are reported in millisecond (ms) and were calculated with a 3-parameter Levenberg-Marquardt curve fitting algorithm. We reported the peak T1 values across the five calf muscle compartments including the anterior (AM), lateral (LM), and deep posterior (DM) muscle groups; soleus (SM), and gastrocnemius (GM) muscles. Averaged peak blood pool T1 values were obtained from the posterior (PT) and anterior tibialis (AT) and peroneal artery (PE). PAD ECV was calculated for each muscle group as (1-hematocrit in %) x [ (1/T1m post) - (1/T1m pre)] / [ (1/T1b post) - (1/T1b pre)], where T1m denotes the maximum T1 value of the respective muscle compartments and T1b the averaged peak arterial blood T1 value.

Variable normality was determined with the Shapiro-Wilk test. Group differences for categorical variables were analyzed with the Chi-square or Fisher exact test. Continuous variables were analyzed with an independent samples Student’s t-test, and the Mann-Whitney-Wilcoxon test was used for non-normal variables. Associations between T1 times and markers of PAD were determined using linear regression. Sante DICOM Editor 3D was used for muscle compartment and lower extremity tracings, T1 extrapolation, and T1 mapping. Intra- and interclass correlations, performed on Stata with the ICC command using DICOM outputs from Sante, were conducted to assess variability in tracing and MRI data analysis between and within operators.

All statistical tests were two-sided, and a *p*-value <0.05 was used as the cutoff for statistically significant phenomena. The statistical analyses were performed with Stata Statistical Software: Release 13 (College Station, Texas, StataCorp LP).

## Results

### Patient Demographics

Patient demographic and clinical characteristics are shown in **Table 1**. Briefly, 66 individuals were enrolled, 5 participants did not complete baseline imaging, an additional 24 participants were excluded due to poor image quality or incomplete MRI exams. A total of 37 participants were included in the analysis (18 PAD patients, 19 matched-controls). Patients were age and gender matched, and there was an equivalent distribution of race among the respective groups. PAD patients compared to controls were more likely hypertensive and hyperlipidemic and had a higher rate of smoking history and prior lower extremity revascularization. As expected, PAD patients had a significantly lower ABI, shorter PWT, and were less likely to complete 6-minutes of treadmill walking compared with the matched-controls.

**Table 1.** Baseline patient characteristics.

| **Variables** | **PAD Patients (n=18)** | **Controls (n=19)** | **P-value** |
| --- | --- | --- | --- |
| Age (years) | 67.6 ± 7.19 | 65.4 ± | 0.43 |
| Males, n (%) | 11 (61.1) | 11 (57.9) | 0.84 |
| Black race, n (%) | 4 (22.2) | 4 (21.1) | 0.86 |
| Body mass index (kg/m<sup>2</sup>) | 27.1 ± 3.85 | 28.2 ± 5.20 | 0.48 |
| Resting ABI | 0.734 ± 0.22 | 1.16 ± 0.087 | <0.0001 |
| Delta ABI | 0.179 ± 0.18 | 0.110 ± 0.097 | 0.16 |
| History of smoking, n (%) | 16 (88.9) | 8 (42.1) | 0.006 |
| Diabetes, n (%) | 7 (38.9) | 3 (15.8) | 0.11 |
| Hypertension, n (%) | 17 (94.4) | 11 (57.8) | 0.010 |
| Hyperlipidemia, n (%) | 17 (94.4) | 11 (57.8) | 0.010 |
| Heart rate (bpm) | 68.6 ± 15.9 | 63.7 ± 6.89 | 0.24 |
| Hematocrit (%) | 38.5 ± 5.73 | 41.2 ± 3.67 | 0.14 |
| eGFR (ml/min/1.73m<sup>2</sup>) | 71.7 ± 21.4 | 78.4 ± 18.0 | 0.36 |
| Anticoagulation, n (%) | 8 (44.4) | 3 (16.7) | 0.057 |
| ACE inhibitor, n (%) | 8 (44.4) | 5 (26.3) | 0.25 |
| Beta blocker, n (%) | 9 (50.0) | 7 (36.8) | 0.42 |
| Claudication onset time (sec) | 112.6 ± 102.8 | N/A | <0.0001 |
| Peak walking time (sec) | 298.2 ± 88.0 | 355.3 ± 20.6 | 0.009 |
| Completed 6 min treadmill walking, n (%) | 11 (64.7) | 18 (94.7) | 0.010 |
| Cholesterol-lowering drug use, n (%) | 15 (83.3) | 8 (42.1) | 0.010 |
| Coronary artery disease, n (%) | 8 (44.4) | 4 (21.1) | 0.13 |
| Lower extremity revascularization history, n (%) | 12 (66.7) | 0 (0.0) | <0.0001 |
| Family history of coronary heart disease, n (%) | 11 (61.1) | 8 (42.1) | 0.25 |

Continuous variables are reported as mean ± standard deviation. PAD: peripheral artery disease, BMI: body mass index, eGFR: estimated glomerular filtration rate, ABI: ankle brachial index. PAD patients: delta ABI, n=17; post-treadmill ABI, n=17; eGFR, n=14; claudication onset time, n=17; peak walking time, n=17; Completed 6 min treadmill, n=17. Matched-controls: heart rate, n=18; hematocrit, n=14; eGFR, n=18; anticoagulation, n=18; coronary artery disease, n=18. N/A in claudication onset time means that there were no claudication in the control group.

### Intra-Observer Reproducibility

Intra-observer reproducibility was excellent for arterial tracings, and the delineations of the leg and the AM, LM, SM, and GM muscle compartments (**Table 2**).

**Table 2.** Intra-observer variability as determined by the intra-class correlation (ICC) coefficient using a two-way model.

| a) |  |  |  |  |  |
| --- | --- | --- | --- | --- | --- |
|  | Intra-observer ICC for the anterior muscle group (right side) | Intra-observer ICC for the lateral muscle group (right side) | Intra-observer ICC for the deep posterior muscle group (right side) | Intra-observer ICC for the soleus muscle (right side) | Intra-observer ICC for the gastrocnemius muscle (right side) |
| Individual ICC (95% CI) | 0.897 (0.794-0.951) | 0.791 (0.594-0.897) | 0.572 (0.274-0.771) | 0.823 (0.658-0.913) | 0.849 (0.707-0.926) |
| Average ICC (95% CI) | 0.946 (0.885-0.975) | 0.883 (0.746-0.946) | 0.728 (0.430-0.871) | 0.903 (0.794-0.954) | 0.919 (0.828-0.962) |
| b) |  |  |  |  |  |
|  | Intra-observer ICC for the anterior muscle group (left side) | Intra-observer ICC for the lateral muscle group (left side) | Intra-observer ICC for the deep posterior muscle group (left side) | Intra-observer ICC for the soleus muscle (left side) | Intra-observer ICC for the gastrocnemius muscle (left side) |
| Individual ICC (95% CI) | 0.914 (0.825-0.959) | 0.875 (0.750-0.939) | 0.707 (0.463-0.851) | 0.924 (0.841-0.964) | 0.805 (0.629-0.903) |
| Average ICC (95% CI) | 0.955 (0.904-0.979) | 0.933 (0.857-0.969) | 0.828 (0.633-0.920) | 0.961 (0.914-0.982) | 0.892 (0.772-0.949) |
| c) |  |  |  |  |  |
|  | Intra-observer ICC for the anterior muscle group (bilateral average) | Intra-observer ICC for the lateral muscle group (bilateral average) | Intra-observer ICC for the deep posterior muscle group (bilateral average) | Intra-observer ICC for the soleus muscle (bilateral average) | Intra-observer ICC for the gastrocnemius muscle (bilateral average) |
| Individual ICC (95% CI) | 0.912 (0.822-0.958) | 0.890 (0.781-0.947) | 0.647 (0.378-0.816) | 0.901 (0.801-0.952) | 0.842 (0.694-0.923) |
| Average ICC (95% CI) | 0.954 (0.902-0.978) | 0.942 (0.877-0.973) | 0.786 (0.548-0.899) | 0.948 (0.889-0.975) | 0.914 (0.819-0.960) |
| d) |  |  |  |  |  |
|  | Intra-observer ICC for the cross-sectional leg area (bilateral average) | Intra-observer ICC for the anterior tibialis artery (right side) | Intra-observer ICC for the anterior tibialis artery (left side) | Intra-observer ICC for the posterior tibialis artery (right side) | Intra-observer ICC for the posterior tibialis artery (left side) |
| Individual ICC (95% CI) | 0.925 (0.847-0.964) | 0.984 (0.984-0.985) | 0.977 (0.975-0.978) | 0.921 (0.918-0.924) | 0.972 (0.970-0.975) |
| Average ICC (95% CI) | 0.961 (0.917-0.982) | 0.992 (0.992-0.992) | 0.988 (0.987-0.989) | 0.959 (0.957-0.960) | 0.986 (0.985-0.987) |
| e) |  |  |  |  |  |
|  | Intra-observer ICC for the peroneal artery (right side) | Intra-observer ICC for the peroneal artery (left side) |  |  |  |
| Individual ICC (95% CI) | 0.876 (0.858-0.891) | 0.914 (0.910-0.917) |  |  |  |
| Average ICC (95% CI) | 0.934 (0.924-0.942) | 0.955 (0.953-0.957) |  |  |  |

ICC and confidence interval were calculated using a two-way model for 26 cases. ICC: intra-class correlation, CI: confidence interval.

### Skeletal Muscle Native T1 Mapping

Native maximum T1 values averaged over the five skeletal muscle compartments (AM, LM, DM, SM, and GM) were significantly elevated in PAD patients compared with matched-controls (1902 (1877-1924) ms vs.1823 (1709-1883) ms, p=0.005; **Table 3**). Native maximum T1 values were significantly higher in PAD patients compared with controls for the AM, LM, DM (all p<0.03) but not for the SM and GM skeletal muscle compartments (Table 3). Native minimum T1 times were not different across the calf muscle compartments.

**Table 3a.** MRI measurements.

| Variables | PAD Patients (n=18) | Controls (n=19) | P-value |
| --- | --- | --- | --- |
| Maximum T1 of composite arteries (ms) | 1729 (1679-1810) | 1688 (1637-1756) | 0.17 |
| Cross-sectional area, anterior muscle group (mm<sup>2</sup>) | 795 (678-980) | 971 (802-1309) | 0.045 |
| Cross-sectional area, lateral muscle group (mm<sup>2</sup>) | 454 (385-554) | 597 (483-803) | 0.007 |
| Cross-sectional area, deep posterior muscle group (mm<sup>2</sup>) | 677 (531-803) | 624 (491-947) | 1.00 |
| Cross-sectional area, soleus muscle (mm<sup>2</sup>) | 1459 (1235-1882) | 1759 (1333-2322) | 0.18 |
| Cross-sectional area, gastrocnemius muscle (mm<sup>2</sup>) | 1326 (1099-1800) | 1649 (1204-2146) | 0.11 |
| Average cross-sectional area (mm<sup>2</sup>) | 954 (828-1175) | 1175 (954-1546) | 0.055 |
| Maximum T1, anterior muscle group (ms) | 1835.5 (188) | 1746 (163) | 0.027 |
| Maximum T1, lateral muscle group (ms) | 1907 (65) | 1755 (279) | 0.002 |
| Maximum T1, deep posterior muscle group (ms) | 1917.5 (106) | 1782 (296) | 0.012 |
| Maximum T1, soleus muscle (ms) | 1930 (143) | 1899 (164) | 0.29 |
| Maximum T1, gastrocnemius muscle (ms) | 1945.5 (50) | 1934 (160) | 0.74 |
| Minimum T1, anterior muscle group (ms) | 878 (777-963) | 878 (777-963) | 0.53 |
| Minimum T1, lateral muscle group (ms) | 897 (818-955) | 840 (718-926) | 0.45 |
| Minimum T1, deep posterior muscle group (ms) | 729 (637-802) | 712 (595-889) | 0.87 |
| Minimum T1, soleus muscle (ms) | 728 (643-813) | 823 (797-912) | 0.024 |
| Minimum T1, gastrocnemius muscle (ms) | 730 (524-900) | 651 (0-809) | 0.55 |
| Average cross-sectional maximum T1 (ms) | 1902 (1877-1924) | 1823 (1709-1883) | 0.005 |
| Average cross-sectional mean T1 (ms) | 1218 (1143-1263) | 1190 (1140-1258) | 0.45 |
| ECV, anterior muscle group (%) | 26.4 (21.2-31.5) | 17.3 (10.2-25.1) | 0.046 |
| ECV, lateral muscle group (%) | 21.7 (15.1-31.2) | 24.7 (19.6-38.5) | 0.43 |
| ECV, deep posterior muscle group (%) | 29.0 (22.5-36.1) | 24.1 (16.5-31.0) | 0.19 |
| ECV, soleus muscle (%) | 22.7 (19.5-27.8) | 13.8 (10.2-19.1) | 0.020 |
| ECV, gastrocnemius muscle (%) | 21.8 (15.1-26.5) | 16.8 (13.3-23.6) | 0.38 |
| ECV, averaged over 5 muscle compartments (%) | 22.9 (21.0-27.5) | 24.5 (20.7-28.0) | 0.68 |

Values are reported as median (interquartile range, IQR); PAD: peripheral artery disease; ms: milliseconds; ECV: extracellular volume fraction. Composite denotes the T1 average for the peroneal, posterior tibialis, and anterior tibialis arteries combined. Average cross-sectional: average T1 times over all five muscle groups. ECV in controls (n=12), ECV in PAD patients (n=16).

**Table 3b.** MRI measurements in diabetic vs. non-diabetic patients with peripheral artery disease.

| Variables | Diabetic PAD<br>Patients (n=7) | Non-Diabetic PAD<br>Patients (n=11) | P-value |
| --- | --- | --- | --- |
| Maximum T1, anterior muscle group (ms) | 1955 + 204 | 1811 + 138 | 0.15 |
| Maximum T1, lateral muscle group (ms) | 1897 + 134 | 1921 + 136 | 0.89 |
| Maximum T1, deep posterior muscle group (ms) | 1903 + 48 | 1916 + 80 | 0.19 |
| Maximum T1, soleus muscle (ms) | 1980 + 250 | 1917 + 127 | 0.26 |
| Maximum T1, gastrocnemius muscle (ms) | 1947 + 125 | 1932 + 50 | 0.62 |
| Average cross-sectional, maximum T1 (ms) | 1924 + 83 | 1911 + 39 | 0.14 |
| Average cross-sectional, mean T1 (ms) | 1270 + 138 | 1227 + 84 | 0.49 |
| ECV, anterior muscle group (%) | 23.2 + 13 | 28.1 + 10 | 0.33 |
| ECV, deep posterior muscle group (%) | 28.5 + 20 | 29.0 + 10 | 0.91 |
| ECV, lateral muscle group (%) | 21.3 + 16 | 23.1 + 16 | 0.66 |
| ECV, soleus muscle (%) | 19.5 + 9.1 | 24.6 + 7.7 | 0.13 |
| ECV, gastrocnemius muscle (%) | 16.8 + 14.9 | 22.7 + 9.1 | 0.33 |
| ECV, average cross-sectional (%) | 26.8 + 8.12 | 22.8 + 2.5 | 0.33 |

PAD: peripheral artery disease; ms: milliseconds; ECV: extracellular volume fraction. Average cross-sectional: average T1 times or ECV over all five muscle groups. Diabetic PAD patients (n=6), Non-diabetic PAD patients (n=10).

### Skeletal Muscle ECV

Skeletal muscle ECV was significantly higher in PAD patients compared with controls for the AM (26.4 (21.2-31.5) % vs. 17.3 (10.2-25.1) %, p=0.046) and SM (22.7 (19.5-27.8) % vs. 13.8 (10.2-19.1) %, p=0.020), but not for the LM, DM, and GM.

### Native maximum T1 and ECV

T1, ECV expression, cross-sectional muscle compartment area were compared among PAD patients and the control group (Table 3a). The composite averaged cross-sectional area of the lower extremity of PAD patients trended lower compared with the controls (*p* = 0.055). The composite maximum T1 for the entire cross-sectional area of the lower extremity was significantly higher in PAD patients versus that in the controls (*p* = 0.005). No significant differences were observed in T1 and ECV values of PAD patients with diabetes compared to those without (Table 3b).

## Discussion

The major finding of this work was that native maximum T1 times averaged over five skeletal muscle groups were higher in PAD patients compared to matched controls. This creates an opportunity for future prospective studies of T1 times and PAD presence and severity.

Prior works have demonstrated the value of T1-mapping in coronary artery disease and left ventricular dysfunction. The findings of this study add to the growing evidence that T1-mapping is of interest in assessing cardiovascular disease. Our study has limitations. The heterogeneity of findings, namely with regard to T1 relaxation times. Additionally, the small sample size of this work precludes generalizability of its findings but does provide early groundwork that future works with larger patient cohorts can expound upon to further assess the role of native peak T1 and ECV in the study of PAD.

In conclusion, native peak T1 values and ECV fractions of several calf muscle compartments are higher in PAD patients compared with matched controls. These data suggest a potential utility of ECV to noninvasively determine skeletal muscle fibrosis in PAD.

## Data Availability

The data supporting the findings of this study are available from the corresponding author upon reasonable request.

## Acknowledgements

The authors wish to thank the participants of this study for their dedication and commitment throughout the work.

## Reference

1. Al-Zaiti S, Besomi L, Bouzid Z, Faramand Z, Frisch S, Martin-Gill C, Gregg R, Saba S, Callaway C and Sejdić E. Machine learning-based prediction of acute coronary syndrome using only the pre-hospital 12-lead electrocardiogram. Nature communications. 2020;11:1–10.

2. Baessler B, Luecke C, Lurz J, Klingel K, Das A, von Roeder M, de Waha-Thiele S, Besler C, Rommel K-P and Maintz D. Cardiac MRI and texture analysis of myocardial T1 and T2 maps in myocarditis with acute versus chronic symptoms of heart failure. Radiology. 2019;292:608–617.

3. Forghani R, Savadjiev P, Chatterjee A, Muthukrishnan N, Reinhold C and Forghani B. Radiomics and artificial intelligence for biomarker and prediction model development in oncology. Computational and structural biotechnology journal. 2019;17:995.

4. Gimnich OA, Singh J, Bismuth J, Shah DJ and Brunner G. Magnetic resonance imaging based modeling of microvascular perfusion in patients with peripheral artery disease. Journal of biomechanics. 2019;93:147–158.

5. Harisinghani MG, O’Shea A and Weissleder R. Advances in clinical MRI technology. Science Translational Medicine. 2019;11:2591.

6. Jellis CL and Kwon DH. Myocardial T1 mapping: modalities and clinical applications. Cardiovascular diagnosis and therapy. 2014;4:126.

7. Ko ES and Morris EA. Abbreviated magnetic resonance imaging for breast cancer screening: concept, early results, and considerations. Korean journal of radiology. 2019;20:533–541.

8. Koçak B, Durmaz ES, Ates E and Kiliçkesmez Ö. Radiomics with artificial intelligence: a practical guide for beginners. Diagnostic and interventional radiology. 2019;25:485.

9. Kuhl CK, Schrading S, Strobel K, Schild HH, Hilgers R-D and Bieling HB. Abbreviated breast magnetic resonance imaging (MRI): first postcontrast subtracted images and maximum-intensity projection—a novel approach to breast cancer screening with MRI. Journal of Clinical Oncology. 2014;32:2304–2310.

10. Lau L, Lo ZJ, Hong Q, Yong E, Zhang L, Chandrasekar S and Tan GWL. LEA 18. Systematic Review on the Use of Artificial Intelligence in Peripheral Vascular Diseases. Journal of Vascular Surgery. 2019;70:122.

11. Mueller T, Hinterreiter F, Luft C, Poelz W, Haltmayer M and Dieplinger B. Mortality rates and mortality predictors in patients with symptomatic peripheral artery disease stratified according to age and diabetes. Journal of vascular surgery. 2014;59:1291–1299.

12. Olin JW and Sealove BA. Peripheral artery disease: current insight into the disease and its diagnosis and management. Mayo Clinic Proceedings. 2010;85:678–692.

13. Ross EG, Shah NH, Dalman RL, Nead KT, Cooke JP and Leeper NJ. The use of machine learning for the identification of peripheral artery disease and future mortality risk. Journal of vascular surgery. 2016;64:1515–1522.

14. Sanchez-Martinez S, Duchateau N, Erdei T, Kunszt G, Aakhus S, Degiovanni A, Marino P, Carluccio E, Piella G and Fraser AG. Machine learning analysis of left ventricular function to characterize heart failure with preserved ejection fraction. Circulation: cardiovascular imaging. 2018;11:e007138.

15. Shieh MC, Belousova T, Taylor AA, Nambi V, Morrisett JD, Ballantyne CM, Bismuth J, Shah DJ and Brunner G. Contrast-Enhanced Magnetic Resonance Imaging Based T1-Mapping and Extracellular Volume Fractions are Associated With Peripheral Artery Disease. Circulation. 2017;136:A17267.

16. Than MP, Pickering JW, Sandoval Y, Shah ASV, Tsanas A, Apple FS, Blankenberg S, Cullen L, Mueller C and Neumann JT. Machine learning to predict the likelihood of acute myocardial infarction. Circulation. 2019;140:899–909.

17. Xiong G, Kola D, Heo R, Elmore K, Cho I and Min JK. Myocardial perfusion analysis in cardiac computed tomography angiographic images at rest. Medical image analysis. 2015;24:77–89.

18. Yang EY, Ghosn MG, Khan MA, Gramze NL, Brunner G, Nabi F, Nambi V, Nagueh SF, Nguyen DT and Graviss EA. Myocardial extracellular volume fraction adds prognostic information beyond myocardial replacement fibrosis. Circulation: Cardiovascular Imaging. 2019;12:e009535.

19. Zhu B, Liu JZ, Cauley SF, Rosen BR and Rosen MS. Image reconstruction by domain-transform manifold learning. Nature. 2018;555:487–492.

